# SIOP Ependymoma I: Final results, long term follow-up and molecular analysis of the trial cohort: A BIOMECA Consortium Study

**DOI:** 10.1101/2021.08.29.21261962

**Authors:** Timothy A Ritzmann, Rebecca J Chapman, John-Paul Kilday, Nicola Thorp, Piergiorgio Modena, Robert A Dineen, Donald Macarthur, Conor Mallucci, Timothy Jaspan, Kristian W Pajtler, Marzia Giagnacovo, Thomas S Jacques, Simon ML Paine, David W Ellison, Eric Bouffet, Richard G Grundy, on behalf of the BIOMECA consortium

**Affiliations:** Children’s Brain Tumour Research Centre, University of Nottingham, UK; Nottingham University Hospitals NHS Trust, UK; Children’s Brain Tumour Research Network (CBTRN), Royal Manchester Children’s Hospital, UK; The Centre for Paediatric, Teenage and Young Adult Cancer, Institute of Cancer Sciences, University of Manchester, UK; The Clatterbridge Cancer Centre, Liverpool, UK; The Christie Hospital Proton Beam Therapy Centre, Manchester, UK; Genetics Unit, ASST Lariana General Hospital, Como, Italy; NIHR Nottingham Biomedical Research Centre; Alder Hey Children’s NHS Foundation Trust; Hopp Children’s Cancer Center Heidelberg (KiTZ), Germany; Division of Pediatric Neuro-oncology, German Cancer Research Center (DKFZ) and German Cancer Consortium (DKTK), Heidelberg, Germany; Department of Pediatric Oncology, Hematology, and Immunology, University Hospital Heidelberg, Germany; UCL GOS Institute of Child Health, London, UK; Great Ormond Street Hospital for Children NHS Foundation Trust, London, UK; Department of Pathology, St. Jude Children’s Research Hospital, Memphis, USA; The Hospital for Sick Children, Toronto, Canada

**Keywords:** Ependymoma, Chemotherapy, Resection, Recurrence, Radiotherapy

## Abstract

**Background:** SIOP Ependymoma I was a non-randomised trial assessing event free and overall survival (EFS/OS) from non-metastatic intracranial ependymoma in children aged 3 to 21 years, treated with a staged management strategy. Chemotherapy efficacy in ependymoma is debated and therefore the response rate (RR) of subtotally resected (STR) ependymoma to vincristine, etoposide and cyclophosphamide (VEC) was an additional primary outcome. We report final results with 12-year follow-up and *post hoc* analyses of recently described biomarkers.

**Methods:** 74 participants were eligible. Children with gross total resection (GTR) received radiotherapy, whilst those with STR received VEC before radiotherapy. DNA methylation and 1q status were evaluated, alongside *hTERT*, ReLA, Tenascin-C, H3K27me3 and pAKT expression.

**Results:** Five- and ten-year EFS was 49.5% and 46.7%, OS was 69.3% and 60.5%. GTR was achieved in 33/74 (44.6%) and associated with improved EFS (p=0.003, HR=2.6, 95% confidence interval (CI) 1.4-5.1). Grade 3 tumours were associated with worse OS (p=0.005, HR=2.8, 95%CI 1.3-5.8). 1q gain and hTERT expression were associated with poorer EFS (p=0.003, HR=2.70, 95%CI 1.49-6.10 and p=0.014, HR=5.8, 95%CI 1.2-28 respectively) and H3K27me3 loss with worse OS (p=0.003, HR=4.6, 95%CI 1.5-13.2). DNA methylation profiles showed expected patterns. 12 participants with STR did not receive chemotherapy; a protocol violation. However, chemotherapy RR was 65.5% (19/29, 95%CI 45.7-82.1), exceeding a prespecified 45% RR.

**Conclusions:** RR of STR to VEC exceeded the pre-specified criterion for efficacy. However, cases of inaccurate treatment stratification highlighted the need for rapid central review. Prognostic associations for 1q gain, H3K27me3 and *hTERT* were confirmed.

## Introduction

Paediatric ependymomas are associated with poor outcomes^1,2^. Five-year overall survival (OS) above 70% is rarely reported whilst event free survival (EFS) is around 55% ^3–6^.

Surgical gross total resection (GTR) is associated with improved outcomes^7–9^. Post-operative radiotherapy of doses up to 59.4Gy using 1.8 Gy per fraction to the tumour bed is now recommended as a standard of care for children over 18 months (54 Gy between 12 and 18 months)^9^. The optimal role for chemotherapy is disputed^10^. Some researchers report benefits of chemotherapy, particularly in younger children^11–15^, whilst others report no benefit, or benefits confined to specific intracranial locations^16,17^.

Progress has been made in understanding the molecular basis of ependymoma. Posterior Fossa A (PFA) and *ZFTA*-fusion (formerly *RELA*-fusion^18^) supratentorial ependymomas are associated with the poorest outcomes^18,19^. Additionally, chromosome 1q gain is a reproducible poor prognostic indicator^1,3,20–23^. Other proposed prognostic markers include telomerase activity via *hTERT*^22,24,25^, Tenascin-C (TNC)^26^, H3K27me3 loss^27,28^, and pAKT expression^29^.

We present previously unreported findings of the International Society of Paediatric Oncology (SIOP) Ependymoma I protocol, recruited between 1999 and 2007, with long-term follow up and retrospective analysis of molecular markers.

The primary aims were:

1. To determine EFS and OS of participants with ependymoma when treated with a staged management strategy to achieve maximum local control;
2. To establish the response rate (RR) of intracranial ependymoma to a combination of vincristine, cyclophosphamide and etoposide (VEC).

## Methods

### Eligibility criteria and outcome measures

Eligible patients were aged 3 to 21 years with untreated non-metastatic, intracranial, histologically confirmed ependymoma. Written informed consent was required. Patients with myxopapillary ependymoma, subependymoma or ependymoblastoma were excluded. Tissue availability for molecular analysis was not mandated for trial entry.

Outcome measures were: EFS and OS, surgical operability and RR to chemotherapy.

### Trial design

Extent of resection was categorised, according to contemporary standards^30,31^, as either subtotal resection (STR) (greater than 1.5cm^2^ tumour residuum on a single cross sectional image) or GTR (no visible residuum, or residuum measuring no greater than 1.5cm^2^) and was determined by radiological consensus, based initially on local opinion before central review. Second look surgery was recommended for those with operable residuum.

Participants with GTR underwent focal radiotherapy of 54 Gy in 30 daily fractions of 1.8Gy per fraction over six weeks.

Participants with STR received chemotherapy with four cycles of VEC with MRI assessments after cycles two and four. Response assessments were centrally reviewed and recorded according to contemporary SIOP recommendations^30^. Percentage response was determined by calculating the product of the perpendicular diameters of the tumour relative to the baseline, post-operative, evaluation.

- Complete Response (CR): no disease;
- Partial Response (PR): 50% reduction;
- Objective Response (OR): 25-50% reduction;
- Stable Disease (SD): <25% reduction;
- Progressive Disease (PD): >25% increase in tumour size.

Following chemotherapy, the protocol specified that all participants receive focal radiotherapy (**Figure 1A**).

**Figure 1:**
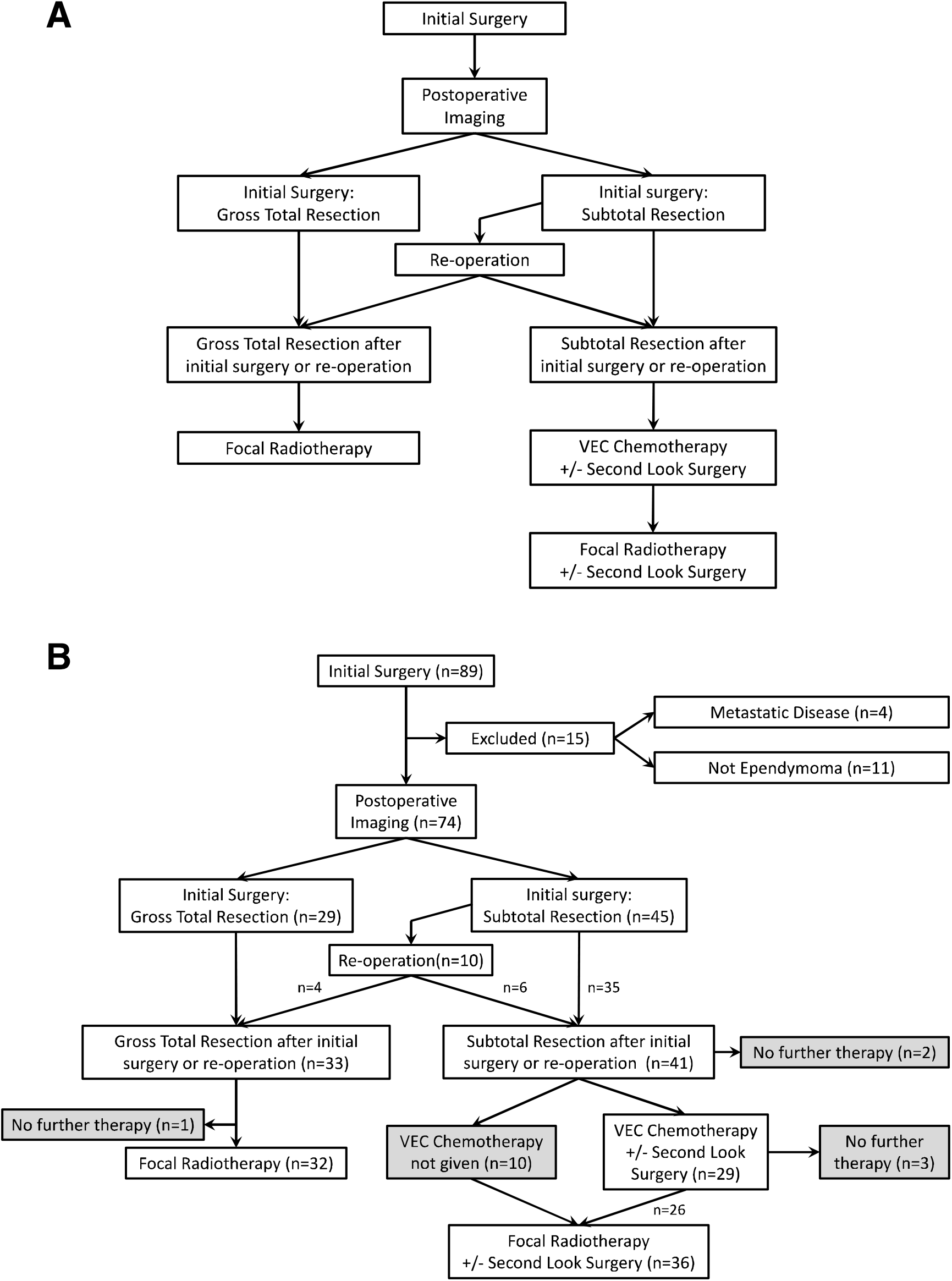
(A) Protocol defined flow through study. (B) Actual flow. 41 had STR and 33 had GTR. Of those with STR, 29 received chemotherapy, 10 received radiotherapy and two had no further therapy. 32/33 with GTR received radiotherapy and one had no further treatment. Grey boxes indicate protocol violations.

The trial was approved by the UK Children’s Cancer Study Group (CSSG) and SIOP. Participants were monitored though the Children’s Cancer and Leukaemia Group (CCLG) data centre until censoring or death. The clinicaltrials.gov identifier was NCT00004224. The study received ethical approval from the Trent multicentre research ethics committee (MREC99/02/11, CTA reference MF8000/13710).

### Molecular Analysis

DNA was extracted from paraffin embedded samples using the⍰AllPrep⍰FFPE DNA/RNA extraction kit (Qiagen) and from frozen samples using the QIAamp DNA mini kit (Qiagen). RNA from frozen samples was extracted using mirVana (ThermoFisher).

DNA methylation profiles were generated using Infinium HumanMethylation450 BeadChip arrays (Illumina) with ependymoma subgroups assigned as previously described^1^. Classifier scores of 0.7 or above were used to support clinical diagnosis, although 31/35 cases had a score >0.9. DNA methylation. IDAT files are accessible at GSE182707.

1q status was evaluated by DNA methylation profiling, fluorescent in-situ hybridisation (FISH) and multiplex ligation-dependent probe amplification (MLPA) as previously described^1^. There was good correlation between 1q assessment methods. The Fleiss-Kappa statistic was 0.615 (p<0.001) and 0.708 (p=0.008) where two and three tests were performed respectively.

Quantitative real-time PCR was used to determine *hTERT* expression as previously described^25^.

Immunohistochemistry was performed in triplicate on four micrometre tissue microarray sections for ReLA, TNC, H3K27Me3 and pAKT. Antigen retrieval was with sodium citrate buffer (pH6). Protein blocking was with normal goat or horse serum (Vector Labs) before five minute peroxidase blocking (DAKO). H3K27me3 (Tri-Methyl-Histone H3, Rabbit mAb, 1:500; Cell Signalling Technology), pAKT (Phospho-Akt, ser473, Rabbit mAb, 1:50; Cell Signalling Technology), and TNC (Tenascin-C, E-9, Mouse mAb, 1:50; Santa Cruz Biotec) primary antibodies were incubated for one hour at room temperature. Positive controls were human tonsil (H3K27me3, RelA), breast carcinoma (pAKT), and epidermoid carcinoma (TNC). Target antigens were detected using the DAKO Envision Detection Kit. For TNC, biotinylated universal antibody anti-rabbit/mouse (Vector, UK) diluted in horse serum was applied, followed with Vectastain avidin/biotin complex reagent (Vectastain-Elite ABC kit, Vector, UK). For all target antigens, diaminobenzidine chromogen was applied for five minutes. H3K27me3, RelA, pAKT and TNC were scored as either negative or positive.

### Statistics

Previous studies indicated five-year OS for those with primary GTR and STR ranges from 30 to 85% and 0 to 45% respectively^32^. This study assumed five-year OS of 70% with GTR and 35% with STR, implying hazard ratio, HR = 0.34 favouring GTR. It was assumed that 3/8 would achieve GTR. Using a two-sided 5% significance level and 80% power, a study of 65 participants was proposed (GTR 24, STR 41)^33^. It was anticipated that if RR to chemotherapy in the STR group was under 25% there would be no interest in the combination. In contrast, RR 45% or more would suggest worthwhile efficacy and was set as the criterion for assessment of chemotherapy response. A one-stage Fleming-A’Hern design required 32 STR participants with a minimum of 13 responses to claim sufficient activity^34^.

### Analysis

EFS was defined as the time from surgery to first recurrence, PD or death. OS was defined as time from surgery to death. Surviving participants were censored at date last seen.

Data analysis was conducted in the R statistical environment^35^. Survival probabilities were calculated using the Kaplan-Meier method^36^. The influences of tumour resection, WHO grade and molecular parameters were investigated prospectively. Molecular analyses were *post hoc*. Multivariate analyses were conducted using the Cox Proportional Hazards Model.

RR was calculated as the proportion of CR plus PR from all those with STR receiving chemotherapy. 95% Confidence Intervals (CI) for the RR were calculated using the Clopper-Pearson approach^37^.

Due to poor compliance with chemotherapy in the STR arm of the study an “as-treated” analysis was performed for chemotherapy treated participants to better understand the true RR to VEC.

## Results

### Cohort Summary

89 participants with intracranial ependymoma from 25 participating centres in the United Kingdom, Ireland, Spain, Denmark, Sweden and the Netherlands were registered between 17^th^ May 1999 and 1^st^ November 2007. Four participants had metastatic disease at presentation. A further 11 did not have ependymoma on histopathology and DNA methylation review (three medulloblastomas, three glioblastomas and one each of atypical teratoid/rhabdoid tumour, choroid plexus carcinoma, high grade glioma, papillary glioneuronal tumour and high grade neuroepithelial tumour with *MN1* alteration). This left 74 with non-metastatic intracranial ependymoma, 35 with a DNA methylation result (**Figures 1B & 2A**).

**Figure 2:**
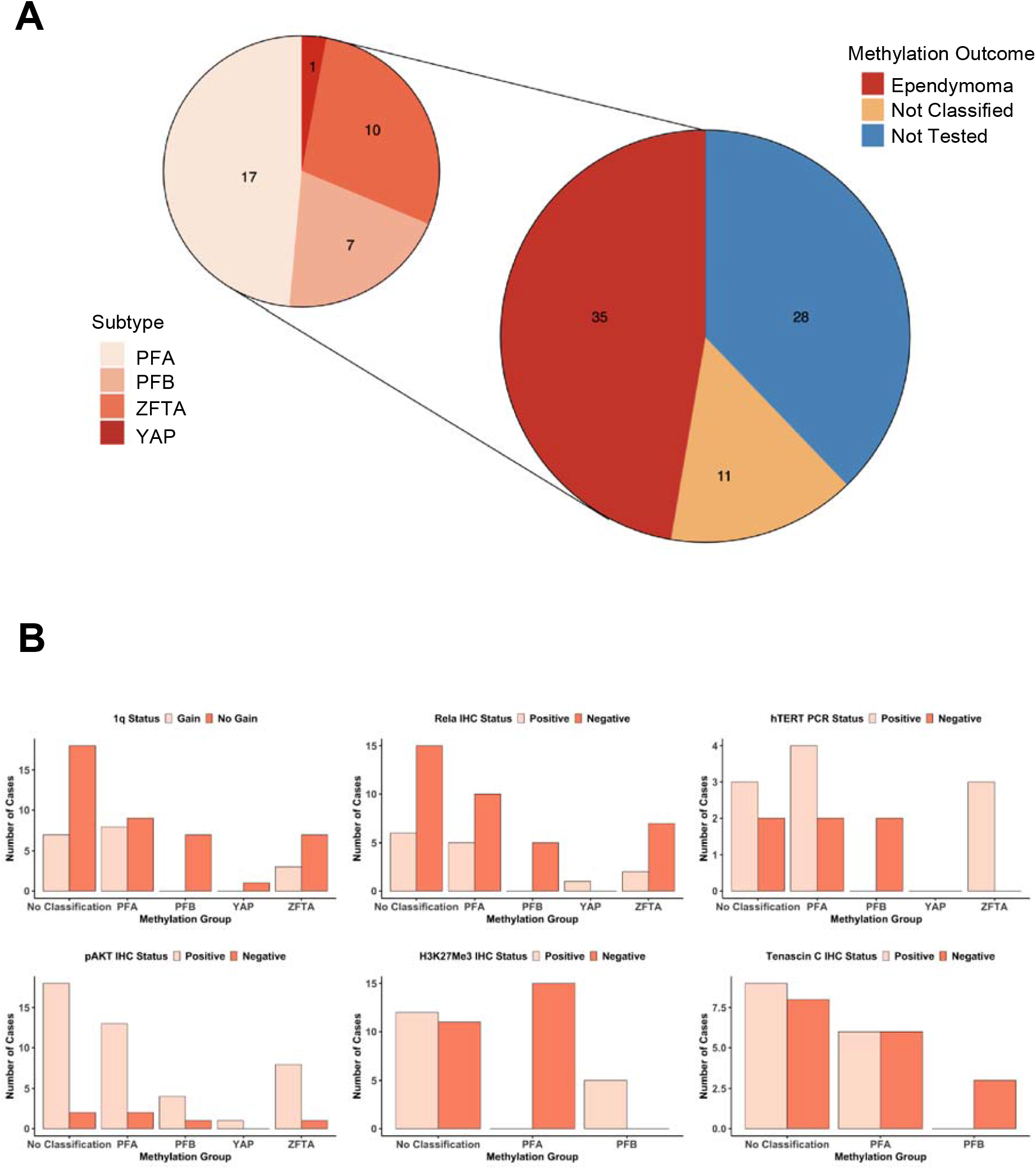
(A) Subdivision of methylation results. 35 participants had a diagnosis of ependymoma assessed by methylation array. Of these 17 were PFA, 10 ZFTA-fusion, 7 PFB and one YAP. Of the remaining 39 cases in the trial, 11 tumours did not have a sufficient classifier score to be assigned and 28 cases had no tissue for analysis. (B) Outcomes of molecular and immunohistochemical analyses stratified by methylation classification. 1q gain seen only in PFA and ZFTA-fusion and hTERT only in PFA and ZFTA-fusion. In posterior fossa tumours, Tenascin-C and H3K27me3 were only identified in PFA and PFB respectively. ReLA IHC was not a good marker for ZFTA tumours as five cases of PFA demonstrated RELA positivity, possibly demonstrating NfkB activation within the tumour microenvironment.

Thirty-eight of seventy-four (51%) participants were male. Median age at diagnosis was 7.8 years (range 3.1-18.8). Forty-seven of seventy-four (64%) had posterior fossa (PF) tumours and 39/74 (53%) were WHO Grade 2. Twenty-nine (39%) participants achieved GTR after first surgery. Of the 45 with initial STR, ten (22%) had early second-look surgery, four of these (40%) achieved GTR and six (60%) still had STR, leaving 41/74 (55.4%) with STR prior to adjuvant therapy and GTR rate of 33/74 (44.6%) (**Figure 1B**). There was no difference in the extent of resection between centres with high numbers of cases (five or more) compared to centres with low numbers of cases (four or fewer) (High volume = 18/39 (46.2%) GTR, Low volume = 15/35 (42.9%) GTR, p=0.776, Chi-square test). There was no difference in age, tumour volume or location between those achieving GTR versus STR. Resection rate improved over time; the GTR rate between 1999-2002 was 12/37 (32.4%), rising to 21/37 (56.8%) between 2003 – 2007 (p=0.035, Chi square test).

To assess second look surgery, a surgical panel retrospectively reviewed post-operative scans following STR to independently consider whether they would have attempted further early resection. There were limitations to availability of complete sets of imaging, but where such review was possible the surgical panel would have attempted an early second resection in 10/25 (40%).

Thirty-five of seventy-four (47%) participants had a DNA methylation result and in 60/74 (81%) 1q status was known. Protein expression was measured in 40/74 (54%) for TNC, 50/74 (68%) for pAKT, 54/74 (73%) for H3K27me3 and 51/74 (69%) for RelA. 16/74 (22%) had *hTERT* expression measured.

Seventeen of thirty-five of DNA methylation profiled tumours were PFA (49%) and 10/35 (29%) were ZFTA-fusion (formerly EPN_RELA^18^). 7/35 (20%) were PFB and 1/35 (3%) were *YAP*. There was no difference in STR between PFA (n=11, 65%) and *ZFTA*-fusion (n=5, 50%) (p=0.730). Eighteen of sixty (30%) had 1q gain (**Table 1**). Twenty-four of sixty were assessed by three methods (FISH, MLPA and methylation array), 16 with two methods and 20 with one method. 1q gain was identified in 8/17 (47%) of PFA and 3/10 (30%) *ZFTA*-fusion. 1q gain was not seen in PFB or *YAP*. There was no difference in 1q gain between GTR and STR cases (p=0.611). *hTERT* was only expressed in PFA and *ZFTA*-fusion tumours. RELA was expressed in all tumour subtypes apart from PFB. In PF tumours, H3K27Me3 was only expressed in PFB. pAKT positivity was seen in all subtypes whilst TNC expression was restricted to PFA (**Figure 2B**).

**Table 1:** Summary of key cohort characteristics, overall and event free survival times. CI: Confidence Interval, GTR: Gross Total Resection, STR: Subtotal Resection, PF: Posterior fossa, ST: Supratentorial.

| Demographic Variable |  | Final Cohort (n=74) |
| --- | --- | --- |
| Resection extent | GTR | 33 |
|  | STR | 41 |
| Gender | Male | 38 |
|  | Female | 36 |
| Age (years) | Median | 7.8 |
|  | Range | 3.1-18.8 |
| Site | PF | 47 |
|  | ST | 27 |
| WHO Grade | 2 | 39 |
|  | 3 | 35 |
| DNA Methylation Analysis | PFA | 17 |
|  | ZFTA | 10 |
|  | PFB | 7 |
|  | YAP | 1 |
| 1q Status | Gain | 18 |
|  | No gain | 42 |
| Follow up (years) | Median | 10.0 |
|  | Range | 0.17-19.00 |
| Follow up survivors (years) | Median | 12.4 |
|  | Range | 1.16-19.00 |
| Event Free Survival (%)<br>95% CI | 5-year | 49.5 |
|  |  | 39.3-62.4 |
|  | 10-year | 46.7 |
|  |  | 36.5-59.7 |
| Overall Survival (%)<br>95% CI | 5-year | 69.3 |
|  |  | 59.4-80.9 |
|  | 10-year | 60.5 |
|  |  | 50.1-73.1 |

### Survival Outcomes (Table 2)

Median follow-up for the surviving participants was twelve years (range 1.2 to 19 years). 32/74 (43%) died: 22/41 (54%) with STR and 10/33 (30%) with GTR. 41/74 (55%) relapsed at least once, 28/41 (66%) with STR and 13/33 (40%) with GTR.

**Table 2:**
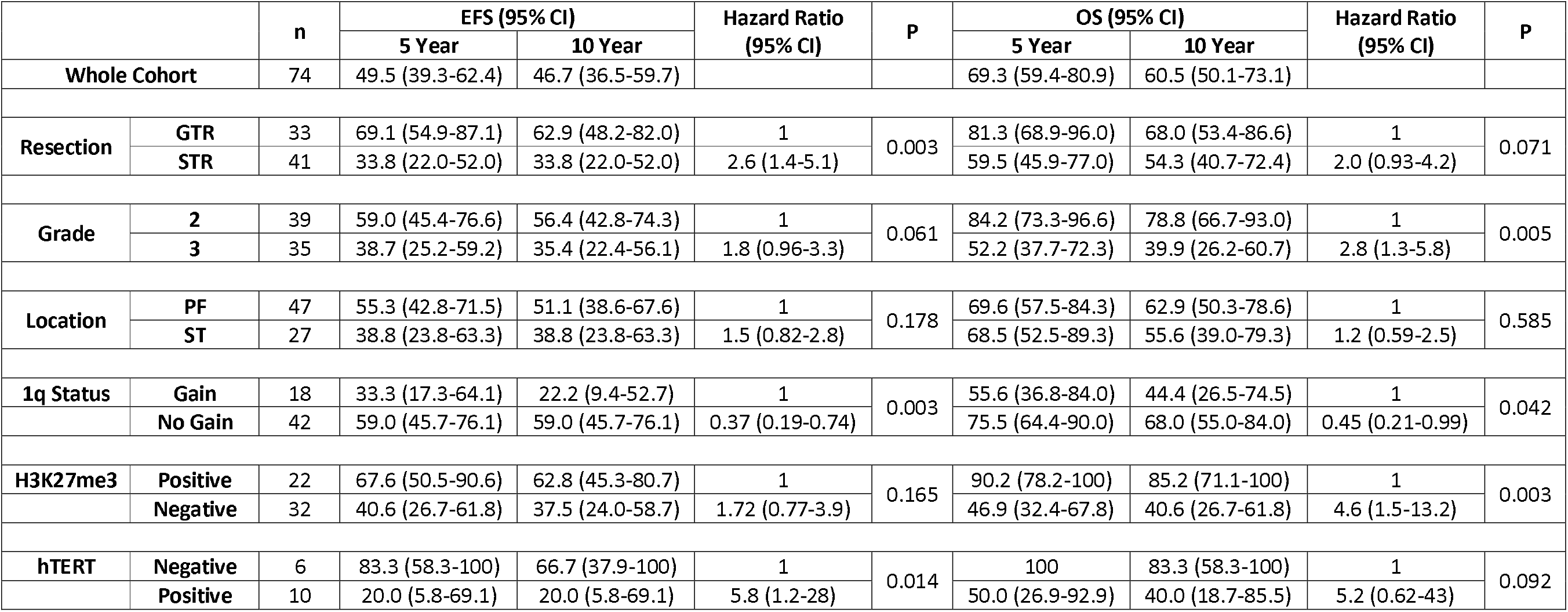
Univariate survival analyses for key clinical and molecular features. Whilst clinical features were defined for analysis prospectively, the additional molecular analyses were *post hoc*. GTR: Gross Total Resection, STR: Subtotal resection, PF: Posterior Fossa, ST: Supratentorial, CI: Confidence Interval, EFS: Event Free Survival, OS: Overall Survival.

Five- and ten-year EFS was 49.5% and 46.7% whilst OS was 69.3% and 60.5%. GTR was associated with improved EFS compared to STR (Five-year EFS 69.1% Vs 33.8%, p=0.003, HR=0.38) (**Figure 3A**) but not OS (Five-year OS 81.3% Vs 59.5.0%, p=0.071, HR=0.50).

**Figure 3:**
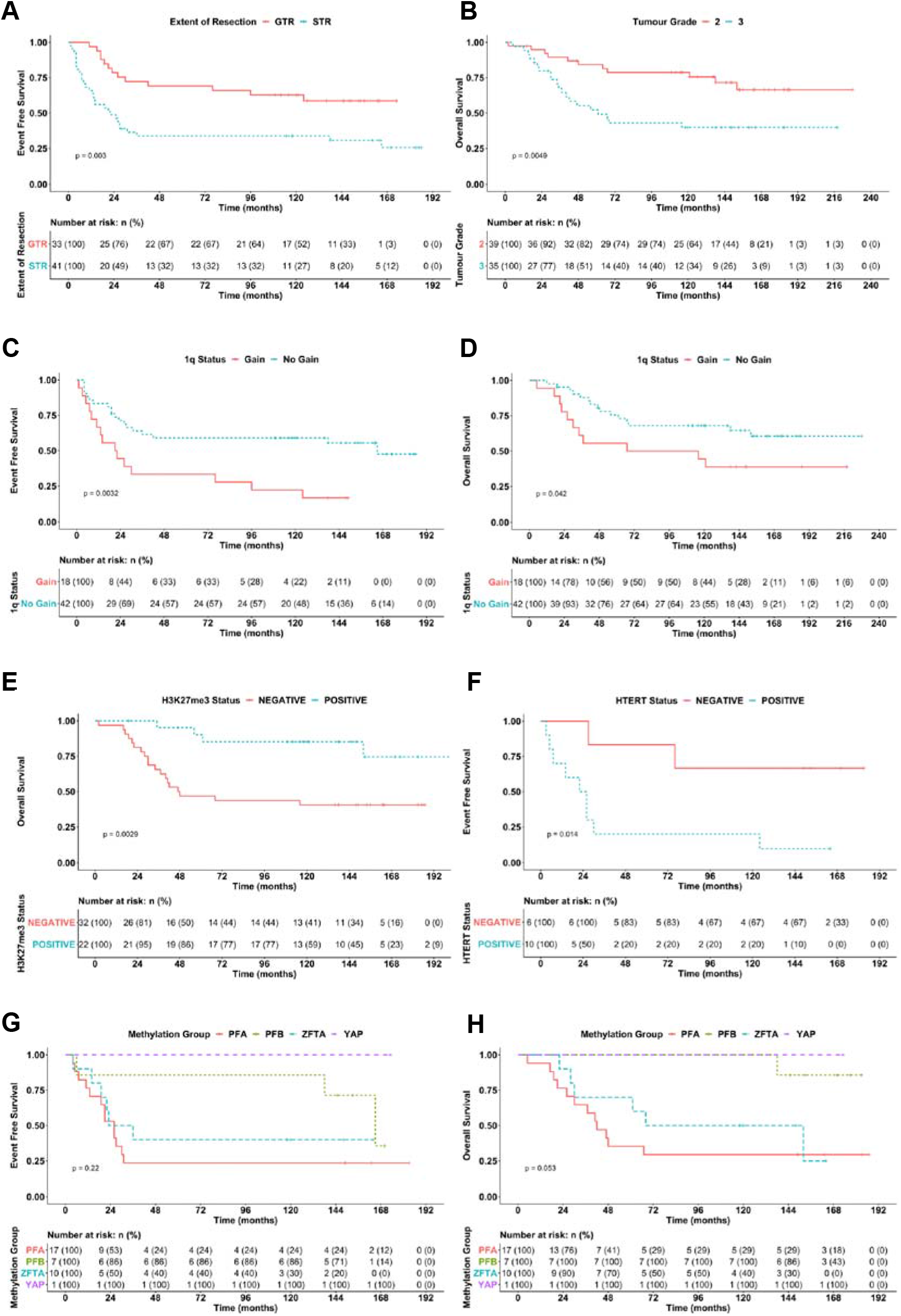
Kaplan-Meier estimators. (A) Participants with STR had poorer EFS than those with GTR. (B) Participant with grade 3 tumours had worse OS than those with grade 2 tumours. Participants with evidence of 1q gain had worse EFS (C) and OS (D). Participants with H3K27me3 loss had worse OS than those with no loss (E). Participants with *hTERT* expression had worse EFS than those with no expression (F). EFS (G) and OS (H) for participants stratified by methylation group did not differ significantly. However, patterns of survival reflect previously published data; lack of significance may relate to low numbers of cases.

Tumour grade was not associated with EFS but WHO grade 3 tumours were associated with worse OS (Five-year OS 52.2% Vs 84.2%, p=0.005, HR=2.8) (**Figure 3B**).

1q gain was associated with poorer EFS (Five-year EFS 33.3% Vs 59.0%, p=0.003, HR=2.71) and OS (Five-year OS 55.6% Vs 75.5%, p=0.042, HR=2.22) (**Figure 3C&D**). When further stratified by tumour location, 1q gain was only prognostic in PF tumours (five-year EFS 33.3% Vs 70.8%, p=0.002, HR=3.94 and OS 58.3% Vs 75%, p=0.023, HR=3.07) (n=36). No difference in survival was seen for supratentorial tumours in relation to 1q gain (n=24) (**Supplementary Figure S1**).

Consistent with its association with loss of expression in PFA, H3K27me3 positivity was associated with better OS (Five year OS 46.9% Vs 90.2%, p=0.003, HR=0.22) but no significant difference in EFS (p=0.200) (**Figure 3E**). *hTERT* positivity was associated with worse EFS (Five-year EFS 20.0% Vs 83.3%, p=0.014, HR=5.8) but not OS (p=0.092) (**Figure 3F**). RELA protein, TNC and pAKT expression were not associated with outcome in any group. DNA Methylation classification was not significantly associated with outcome, possibly due to small numbers of cases, but patterns of survival were consistent with previous reports^1,19^ (**Figures 3G&H**).

On multivariate analysis of OS including 1q status and grade, only grade 3 tumours remained a predictor of poorer outcome (p=0.015, HR=2.81, 95%CI 1.22-6.47). For EFS, both 1q status and resection remained statistically significant (1q gain: p=0.003, HR=2.87, 95%CI 1.43-5.78, STR: p=0.006, HR=2.81, 95%CI 1.35-5.84).

### Treatment outcomes

In the GTR group (n=33), 32 participants (97%) received focal radiotherapy. One participant, with supratentorial ependymoma lacking 1q gain, received no further treatment due to family choice and was still alive after ten-years (**Figure 1B**).

In the STR group (n=41), despite a clear protocol, 10 participants with residuum proceeded to radiotherapy without chemotherapy (**Figure 1B**). Of these, four resulted from family preference, and one was because resection was assessed locally as GTR, but STR on subsequent central review. No reason was documented for the remaining five. Two participants received no therapy after STR; one dying within two months of diagnosis and the other progressing after 14 months.

26 STR group participants received chemotherapy followed by focal radiotherapy whilst a further three received chemotherapy alone. Of the 29 who received chemotherapy: 22 received four cycles of VEC; four received two cycles and proceeded to further treatment with progressive disease as per protocol; and three received two or three cycles but did not progress to four cycles due to toxicity.

The CR+PR response rates were 14/28 (50.0%) and 13/22 (59.1%) after VEC cycles two and four respectively. 19/29 (65.5%, 95% CI 45.7-82.1) participants achieved CR or PR during chemotherapy, exceeding the prespecified 45% RR criterion (**Table 3**).

**Table 3:** Summary of responses to chemotherapy as assessed by MR imaging after 2- and 4-cycles and best response achieved within first four cycles of chemotherapy.

| VEC Response |  | 2-Cycle Response | 4-Cycle response | Best Response |
| --- | --- | --- | --- | --- |
| Complete Response | CR | 6 | 5 | 9 |
| Partial Response | PR | 8 | 8 | 10 |
| Objective Response | OR | 2 | 0 | 0 |
| Stable Disease | SD | 6 | 3 | 4 |
| Progressive Disease | PD | 6 | 6 | 6 |
| CR+PR (%) |  | 14/28 (50.0) | 13/22 (59.1) | 19/29 (65.5) |
| 95% CI |  | 30.7-69.4 | 36.4-79.3 | 45.7-82.1 |

In a *post hoc* analysis, the twelve participants in the STR group who did not receive chemotherapy had median OS of 64 months, whilst those who received chemotherapy did not fall below 50% survival during follow-up. However, there was no difference between the two groups for either OS (p=0.35) or EFS (p=0.56).

The main toxicities of chemotherapy were leucopenia (97%), thrombocytopenia (48%), nausea and vomiting (24%), and infection (21%).

Most second look surgeries (10/16, 63%) occurred prior to adjuvant therapy, but because later surgical decisions were taken at local centres, it has not been possible to determine detailed outcomes of second surgery for the remaining six participants.

## Discussion

Whilst knowledge of the molecular basis of ependymoma has advanced since the trial was designed^18,19,38,39^, our ability to treat ependymoma, in particular to prevent recurrence^1^, has shown less progress. We report long-term outcomes for children treated with a staged treatment protocol for ependymoma and retrospectively apply molecular diagnostics to a clinical trial cohort^3,21,25^. We also report that chemotherapy meets a prespecified RR, confirming VEC as standard of care for inoperable childhood ependymoma.

A primary aim of this study was to assess the role of chemotherapy in inducing a tumour response for participants with STR. The RR of 65.5% suggests that VEC is associated with tumour response in this group. Whilst the number receiving chemotherapy was 29 rather than the targeted 32, the 19 responses exceeded the 13 prespecified for demonstration of efficacy. The study was designed to assess RR rather than survival as an outcome, and further work is required to establish whether chemotherapy provides either a direct survival benefit or an indirect benefit through the facilitation of additional surgery. The *post hoc* comparison between STR participants with and without chemotherapy was not designed to answer this question.

In 12 participants for whom STR was achieved, no chemotherapy was given, in breach of the protocol. This resulted from treatment decisions by local teams, as prospective central radiological review for stratification was not mandated. Additionally, there were four cases in which the family did not consent to chemotherapy. The challenge of protocol compliance for post-operative chemotherapy regimens in ependymoma is not a unique experience; a similar problem has been reported in ACNS0831^40^ and the SIOP ependymoma II study is experiencing related difficulties (Personal Communication, R Grundy, 2021). We have included rapid central review within the current SIOP Ependymoma II study and based on our experience, we recommend the use of multidisciplinary meetings at trial registration in order to ensure correct stratification to treatment arms.

Whilst a number of studies have identified a role for chemotherapy in children under three years, aiming to avoid or at least delay radiotherapy^12,13,15,21^, others have disagreed^4,6,14^. Our study demonstrated chemotherapy efficacy in older age groups. This is consistent with the findings of more recent US studies^6,41^. Garvin used a combination of vincristine, cisplatin and etoposide and reported a RR of 57% in 35 evaluable participants, which is close to our RR of 65.5%^6^. Massimino reported a cohort of 160 children using VEC chemotherapy to bridge the gap to second resection, but did not directly report on the RR as assessed by post-chemotherapy imaging^4^. More recently, the ACNS0121 study investigated children with STR and vincristine, carboplatin, cyclophosphamide and oral etoposide, followed by radiotherapy and second look surgery with combined CR and PR of 67%^41^. Additionally, recent results from the ACNS0831 study indicate a role for chemotherapy in some patients with totally, or near totally, resected ependymoma^40^. Our results add to the body of evidence advocating the post-surgical use of chemotherapy prior to further therapy in children with STR. The VEC chemotherapy regimen in SIOP ependymoma I did not include a platinum based drug. Given the evidence of response of tumour residuum to VEC, future studies need to consider the benefits of platinum chemotherapy against the risk of nephrotoxicity and ototoxicity. Unfortunately, SIOP ependymoma I was not designed to answer this important research question.

Although not a primary aim, it was hoped that this trial would answer questions related to surgery. The importance of GTR in obtaining good outcomes was previously known and has been emphasised again in this study, with improved EFS. This study was powered for a 38% GTR rate and bettered this with 45% of participants having GTR prior to adjuvant therapy. At the end of the trial, a retrospective review of scans attempted to elucidate factors behind complete or incomplete surgery. This was hampered by limited availability of complete sets of pre- and postoperative scans and access to operative notes. However, as reported, no differences between GTR or STR were identified.

Trial participation can influence practice and it is notable that GTR rates rose from 32.4% to 56.8% between the first and second halves of the trial. In the years since trial inception, surgeons have become increasingly aware of the prognostic impact of GTR on outcomes for ependymoma. However, they are frequently not aware of the diagnosis at the time of operation. Review of a subgroup of operation notes in this study revealed that smear or frozen section results were frequently uninformative in determining tumour type, being non-committal or incorrect. In less than half of cases in which operation notes were reviewed was there either confident suggestion of ependymoma or confirmed pre-operative histology. Improvements in imaging techniques and magnetic resonance spectroscopy may help to address this challenge in the future^42^.

The small number of participants undergoing further early resection makes it hard to assess the added value of attempting to convert STR to GTR with a second operation. The retrospective surgical panel felt that twice as many participants may have been suitable for second look surgery as actually occurred (40% versus 22%). As a result of this experience, earlier consideration of further surgery by an independent panel alongside close assessment of the morbidity associated with repeat surgery is a focus of the current SIOP Ependymoma II trial^43^. A weekly panel review aims to deliver a more definitive answer on the benefits and morbidity of exhaustively pursuing GTR with repeated surgeries. This is critical, as surgery for cerebellopontine angle ependymoma is associated with high morbidity^44^.

The protocol requested radiotherapy information including: baseline imaging used to determine gross tumour volume; copies of the radiotherapy plan and treatment chart; and copies of the simulator and machine verification films for quality assurance. However, there was no established process in place to facilitate this and hence full radiotherapy dosimetric data was not available for analysis. The trial was carried out during a time of transition from 2D simulator planning, to CT assisted 3D planning and the technical details reflect this. In the 20 years since the design of this study, intensity-modulated radiation therapy, volumetric modulated arc therapy, helical tomotherapy and proton beam therapy have become standard of care for ependymoma and the radiotherapy techniques in this trial do not reflect contemporary practice. Additionally, there has been progress in image guidance and prospective trial quality assurance of both contours and plan parameters.

The prospective clinical trial was designed prior to recent molecular discoveries; however, it provided an opportunity to apply molecular analyses to a well-defined cohort to allow comparison with other ependymoma studies^45^. The results of central histopathological review and DNA methylation classification aligned closely with one another, however around half of the cohort did not undergo DNA methylation profiling. Future studies may mandate the provision of tissue for molecular analyses, because they have the potential to provide more objective tumour classification for prognostication and therapeutic stratification; histopathological grading of ependymomas being subjective and in many studies not a robust prognostic indicator^46^.

The finding that 1q gain was associated with poor outcome confirms previous reports^22,24,25^. In line with ACNS0121 we assessed 1q gain across supratentorial and PF tumours and found it to only be associated with poorer outcomes in children with PF tumours^41^. The prevalence of 1q gain in our cohort was higher than reported by others^20,47,48^. One possibility is that this is a result of inclusion of multiple methods of testing, detecting additional cases that may not have been identified by FISH alone. Alternatively, there may be a genuinely higher rate of 1q gain in our cohort, but we were unable to identify any abnormality that would make this more likely.

*hTERT* mRNA expression was associated with poorer outcomes. However, this analysis was conducted in a small subgroup of the whole cohort. Notwithstanding this, our finding is consistent with the previous reports for ependymoma expressing *hTERT* ^22^. We also confirmed previous findings limiting *hTERT* expression to PFA and *ZFTA*-fusion^23,26^.

Loss of H3K27me3 expression was associated with poorer OS, likely reflecting the strong association between loss of this marker and PFA tumours^27,28^. TNC expression and absence of H3K27me3 was limited to PFA^19^. Given that TNC is a marker for PFA, based on the other samples tested with no DNA methylation result, there was likely to be at least a further nine PFA tumours in our cohort, indicating that 70% of our tumour cohort comprised of PFA. This is consistent with PFA being the most common childhood ependymoma subtype^26,49^. RELA protein has also been reported as an immunohistochemical marker for ZFTA-fusion^26,49^, however, we identified RELA expression across multiple subtypes, indicating that RELA expression alone is not suitable for identifying *ZFTA-*fusion tumours.

The retrospective nature of the methylation analysis resulted in low numbers of each subtype, highlighting the challenges of obtaining sufficient tissue and the risk of the sample either being inadequate or unclassifiable. When designing prospective studies with molecular stratification estimations of cohort sizes must be adjusted to account for this. It is also important to continue to identify reliable but robust markers for molecular subgroups which could be used in place of DNA methylation profiling^50^.

## Conclusions

We present evidence that amongst children with STR there is activity of VEC with a RR of 65.5% (95% CI 45.7-82.1) with an acceptable toxicity profile, supporting a potential beneficial role for chemotherapy in children with intracranial ependymoma. However, as this study was not designed to measure changes in EFS and OS, further data is required to determine whether there is an associated impact on survival. Further work is needed to establish whether this varies depending on the molecular designation of individual tumours. We confirm that 1q gain, *hTERT* expression and loss of H3K27me3 are poor prognostic factors for intracranial ependymoma. Future trials need to include prospective, molecularly stratified, approaches to better understand the clinical implications of recent molecular discoveries.

## Data Availability

Data is related to individual patients but anonymised data can be provided by the authors following a reasonable request.
DNA methylation profile data has been deposited for public access at the gene expression omnibus: GSE182707.

## Acknowledgements

The authors acknowledge the CRUK clinical trials unit team including Andrew Raxworthy-Cooper for the support of this study. TR acknowledges the continued support of ‘Fighting Ependymoma’.

