## Supplementary Figure S1 for "SIOP Ependymoma I: Final results, long term follow-up and molecular analysis of the trial cohort: A BIOMECA Consortium Study"

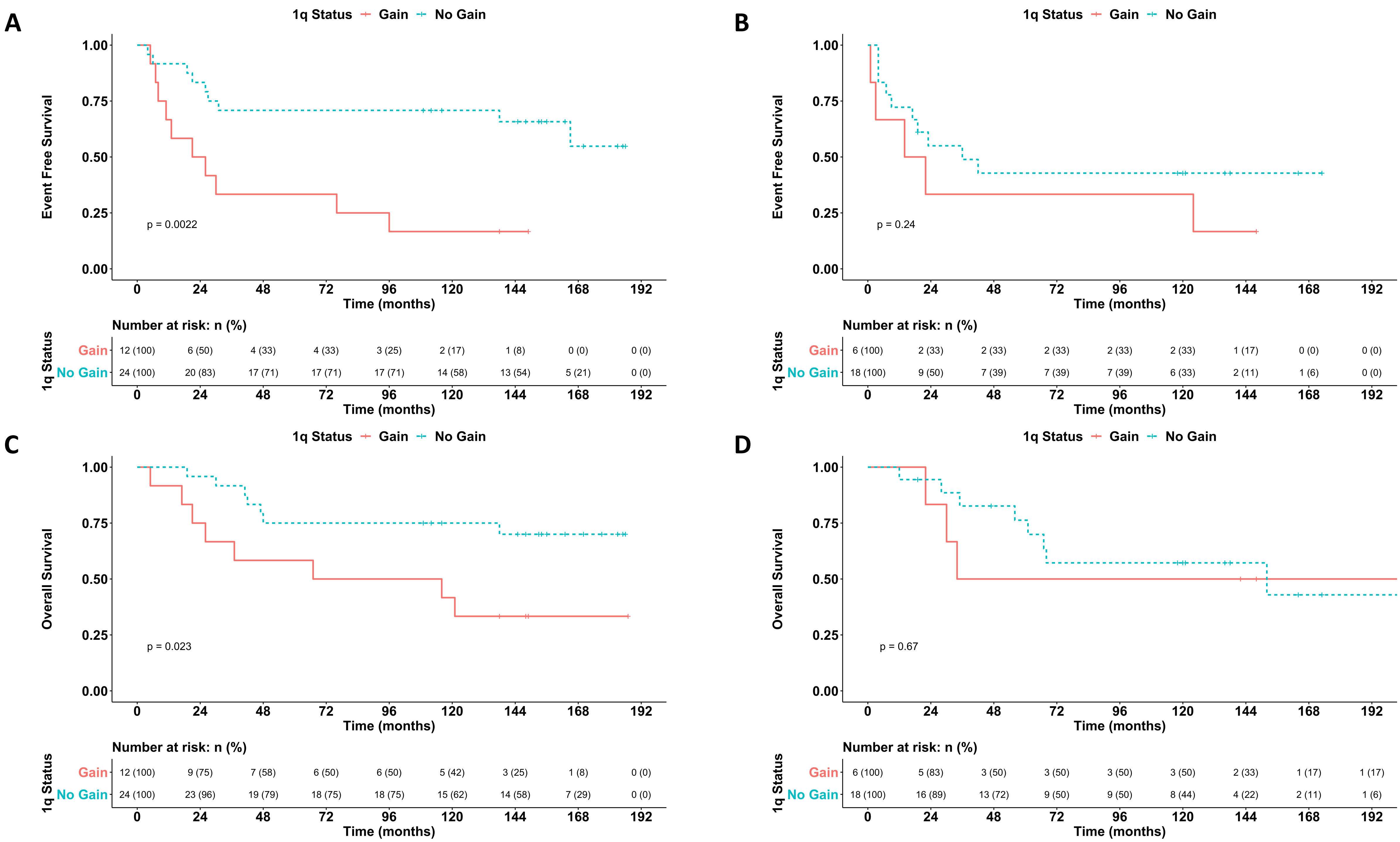


**Supplementary Figure S1:** Association between 1q gain and (A) EFS in posterior fossa tumours, (B) EFS in supratentorial tumours, (C) OS in posterior fossa tumours and (D) OS in supratentorial tumours. Note that the association between outcome and 1q gain is only seen in the posterior fossa cohorts for both survival metrics.
